# Development and validation of 2D-LiDAR-based gait analysis instrument and algorithm

**DOI:** 10.1101/2020.08.21.20178996

**Authors:** Seongjun Yoon, Hee-Won Jung, Heeyoune Jung, Keewon Kim, Suk Koo Hong, Hyunchul Roh, Byung-Mo Oh

**Affiliations:** Dyphi Research Institute, Dyphi Inc., Daejeon 34068, South Korea; Department of Internal Medicine, Seoul National University Hospital, Seoul 03080, South Korea; Department of Rehabilitation Medicine, National Traffic Injury Rehabilitation Hospital, Gyeonggi-do 12564, South Korea; Department of Rehabilitation Medicine, Seoul National University Hospital, Seoul National University College of Medicine, Seoul 03080, South Korea

## Abstract

Acquiring gait parameters from usual walking is important to predict clinical outcomes, including life expectancy, risk of fall, and neurocognitive performance, in older people. For comprehensive gait analysis, instruments such as marker-based motion analysis systems and walkways with pressure sensor arrays are necessary. Traditional instruments are bulky, complex, expensive, and intrinsically intrusive. Requirements of dedicated spaces for installation and specialized staff make it difficult to utilize traditional gait analysis instruments in most outpatient clinics. We present a novel gait analysis tool that is small, highly accurate, easy-to-use, and non-intrusive and is based on two-dimensional light detection and ranging technology. Using an object-tracking algorithm, we conducted a validation study of spatiotemporal tracking of ankle locations of subjects by comparing our tool with a gold standard modality. Our tool showed successful acquisition of gait parameters from usual walking motions and trackability with multiple targets in noisy conditions, including typical clinical environments.

## Introduction

The clinical importance of assessing physical performance is increasing with the rise in the older population globally [1, 2]. In older people, a spectrum of physical performance has been studied for its importance in numerous aspects such as biomarkers of clinical outcome prediction [3], criteria for selecting population eligible for certain intervention programs [4], and outcome measures per se [5]. Given the clinical relevance, items of physical performance are considered key components in assessing common geriatric syndromes, including frailty and sarcopenia [1].

Besides various factors of physical performance such as muscle strength, muscle power, and balance, gait parameters have also been extensively studied [6, 7]. For example, usual gait speed has been studied for its association with life expectancy [8], risk of fall [9], and risk of adverse outcomes after various clinical procedures [10]. Furthermore, other gait parameters such as cadence, step length, and step width have been also shown to be associated with clinical features including neurocognitive performance [11], extents of vascular aging, and other geriatric parameters [12, 13].

However, acquiring gait parameters other than usual gait speed that can be measured by a stopwatch has been a challenging task in both clinical practice and research focusing on older adults. Previously, to measure step length, step width, and cadence, subjects were required to walk through walkways with pressure sensor arrays [14] or performing marker-based motion capture studies [15], both of which are not highly accessible in most geriatric practices even in developed countries. Furthermore, inviting people to facilities with these instruments might be not a feasible option since many geriatric populations reside in long-term care facilities or communities with decreased mobility. Therefore, simple, portable sensor-based gait analysis protocols may improve difficulties in studying gait parameters in the older frail population.

For measuring usual gait speed, we developed sensor-based instruments and showed cross-correlations between modalities including 1D light detection and ranging (LiDAR), infrared, ultrasound, and laser sensors. In this study, to further include gait parameters of step length, step width, and cadence, we developed and validated an instrument and algorithm using the 2D-LiDAR sensor with 3D marker-based motion capture study as a ground truth.

## Methods

### Test environment & participants

To validate the gait monitoring instrument based on the 2D-LiDAR sensor, all tests were performed at a dedicated gait analysis laboratory at National Traffic Injury Rehabilitation Hospital, South Korea. A preinstalled motion capture system was calibrated to collect kinematic data of the participants. Though the entire region of interest (ROI) for motion capture system was 7000 × 2000 mm^2^ (see Fig. 1a), in this study, the area of 6000 × 2000 mm^2^ in the middle of the ROI was mainly monitored to ensure reliable data acquisition from the motion capture system as well as 2D-LiDAR sensor. In addition, a stereo camera, which simulates human binocular vision to estimate spatial depth information from stereo images, was included as a possible alternative to the 2D-LiDAR sensor.

**Figure 1.**
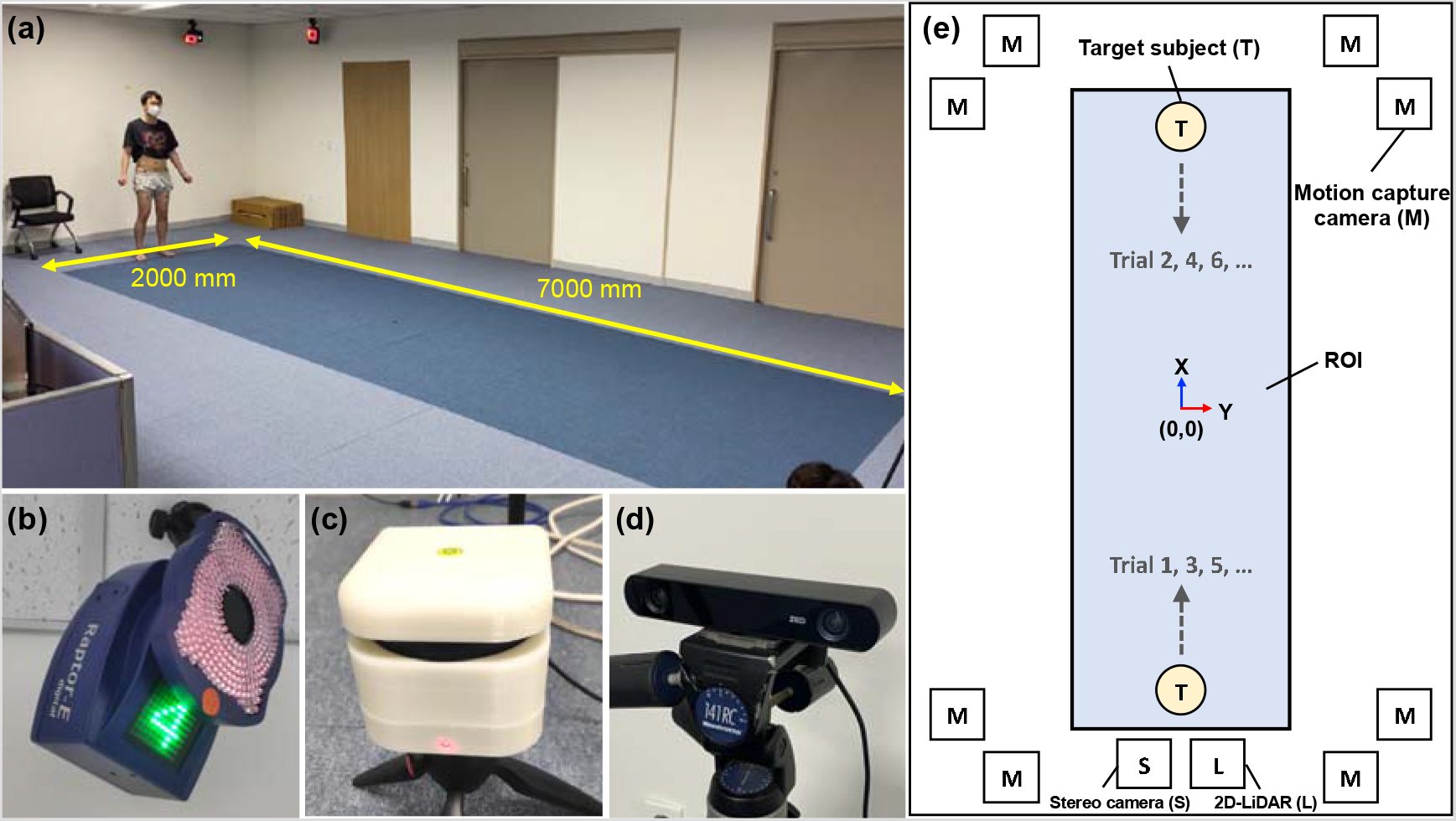
Test environment and instrument settings for validation of 2D-LiDAR-based gait analysis. **a** Gait analysis room showing ROI area of motion capture system marked as blue on the floor. **b** One of eight near infrared cameras of motion capture system. **c** 2D-LiDAR sensor. **d** Stereo camera. **e** A schematic of a test environment with spatial coordinates based on motion capture system. The center of ROI is denoted as (0 mm, 0 mm). The sensor positions of 2D-LiDAR and stereo camera are shown simultaneously.

For the tests, four healthy participants were enrolled (male, aged 30–45). The participants were guided to walk through the forward and reverse directions along the x-axis in each trial (see the walking directions in Fig. 1e and Supplementary Video 1). A total of 40 trials, 10 trials per participant, were conducted. The institutional review board of National Traffic Injury Rehabilitation Hospital approved the study protocol (No. NTRH-20004), and written informed consent was acquired from all participants.

### Instrumentation

The gait data from the motion capture system (Raptor-E, Motion Analysis, USA) were regarded as a ground truth in this study. The gait analysis data from the 2D-LiDAR sensor and stereo camera were evaluated based on the data from the motion capture system. The motion capture system consists of eight near-infrared (750 nm) cameras with a frame size of 1.3 megapixels and a resolution of 1280 × 1024 pixels (Figs. 1a, b). The frame rate was set to 60 frames per second (fps). A marker set consisting of 15 markers was placed at the lower body of the subjects to determine the joint positions during gait analysis (Supplementary Fig. 1). In particular, the markers of left and right ankles were selected for comparison with the spatiotemporal locations obtained from the other sensors.

RPLiDAR A3M1 (Shanghai Slamtec Co., Ltd., China) with a scan rate of 10–15 Hz and an angular resolution of 0.225° was used as the 2D-LiDAR sensor (Fig. 1c, the sensor is enclosed in a self-designed outer package). The maximum distance ranges from sensor specification were 25 m and 10 m for white and dark objects, respectively. In our study, however, typical sensing ranges were limited to 10 m or closer for a reliable detection, accounting for decreased spatial resolution at longer distance. The height of the sensor position was adjusted to detect near ankles of target subjects.

ZED2 (Stereolabs Inc.) was used as the stereo camera (Fig. 1d), which captures high-definition binocular images with a wide field of view (FoV) and generates two synchronized left and right video streams. It estimates depth by stereo-matching method between two scenes from left and right cameras. The FoV was 110° (horizontal) × 70° (vertical) × 120° (depth) and the frame rate was set to 30 fps.

### Object tracking algorithm for 2D-LiDAR sensor

In this study, a novel object tracking algorithm, i.e., the inertia-based object tracking algorithm (IOTA), was developed to recognize left and right ankles from raw pointcloud data of the 2D-LiDAR sensor and to track spatiotemporal locations of the ankles. IOTA is based on the assumption that the velocity of slowly moving objects is negligibly changed if data scan rates are fast [16]. A detail description about IOTA is as follows:

1. A set of pointcloud data is gathered by a single 2D-LiDAR scan, denoted as frame.
2. Initially, an arbitrary point from the pointcloud data is selected. A numeric label is tagged to the initial point, and subsequently, a nearest neighbor point is found. The same numeric label is tagged to the nearest neighbor point if the spatial distance is closer than the threshold. Otherwise, the point is tagged with a new label with a higher number. This is repeated until all points in the set of pointcloud are tagged, and consequently, a set of tagged point groups is generated.
3. An arbitrary tagged point group is selected, and subsequently, the nearest tagged point group is found. The nearest point group is merged to the previously selected point group if the nearest spatial distance between any two points belonging to each point group is closer than the threshold. This is repeated until all the tagged point groups are checked.
4. If the numbers of points in the tagged point groups are smaller than the threshold, the labels of the tagged point groups are dropped, i.e., very small point groups are eliminated.
5. Each tagged point group is regarded as an individual object snapshot. The center of mass (CoM), object size, and curvature are calculated. If the object size and curvature fail to meet the predefined criteria, the object snapshot is dropped.
6. Euclidean distance score is calculated for each object snapshot in the current frame by comparing the change in CoM displacement in inter-frame time, i.e., object velocity, to every object snapshot captured in the previous frame. Here, it is assumed that the time difference between the frames is sufficiently short. For the 2D-LiDAR sensor used in our study, the time difference between two subsequent frames was around 80 ms, which was deemed to be sufficiently fast to analyze typical movements of persons. Therefore, it could be assumed that the velocity change in an object snapshot is negligible between two subsequent frames if the object snapshot originated from the same target, causing the Euclidean distance score to be close to 1.
7. Each object snapshot in the current frame is matched to the most probable object ID, which is predetermined in the previous frames. If there are any contradictions in the current object snapshots during the matching of object ID, for instance, if some object IDs are matched to more than two object snapshots in the current frame, the object ID of the snapshot with the lower Euclidean distance score is rejected and the next probable object ID is selected.
8. If object snapshots are in a starting area, which is predetermined before the measurement, the object IDs are set as targets. The targeted object IDs are assigned to left or right ankles based on the direction of orthogonal vector calculated by velocity vector and counterpart pointing vector. If the number of targeted object snapshots is greater than two, only two object snapshots nearest to the 2D-LiDAR are chosen.
9. If the left and right ankles are targeted, steps 1 to 7 are repeated in every frame. Otherwise, the steps 1 to 8 are repeated in the next frame.

### Object tracking algorithm for stereo camera

For the stereo camera, the spatiotemporal locations of left and right ankles were tracked by combining the 2D human pose estimation (2D-HPE) network with depth data gathered from the stereo camera. The 2D-HPE network was applied to the left camera image for each frame. In this study, a well-trained 2D-HPE network provided by openVINO toolkit (Intel Corp.) was selected [17]. Although the estimated pose contained up to 18 key points, only the key points of left and right ankles were used. The representative pixels of the left and right ankles were converted to 3D points by depth maps corresponding to the pixel maps.

## Results

### Spatiotemporal tracking of ankle locations

For three different modalities of motion capture system, 2D-LiDAR with IOTA, and stereo camera with 2D-HPE, the tracked spatiotemporal locations of ankles along the axial (x-axis) and lateral (y-axis) directions of walking are shown in Figs 2a and b, respectively. The highest frame rate of the motion capture system (60 fps) produced rich tracked points, while low frame rates of 2D-LiDAR sensor (10–15 fps) and stereo camera (30 fps) produced sparse tracked points. In addition, tracking failures, mostly caused by occlusions and far distance from the sensors, further deteriorated the sparsity of tracked points for both of 2D-LiDAR with IOTA and stereo camera with 2D-HPE. Compared to the measured spatial locations of ankles along the axial direction (Fig. 2a), the spatial locations along the lateral direction showed poor agreement with those of the motion capture system for both of 2DLiDAR with IOTA and stereo camera with 2D-HPE (Fig. 2b), because of limited spatial resolutions for each sensor. Slight mismatches between the walking axis (i.e., y = 0), and the sensor positions (see Fig. 1e) also affected the spatiotemporal accuracy especially of the stereo camera with 2D-HPE, where a significant spatial drift was observed, while 2D-LiDAR with IOTA is much tolerable to the spatial drift, as shown in Fig. 2b.

**Figure 2.**
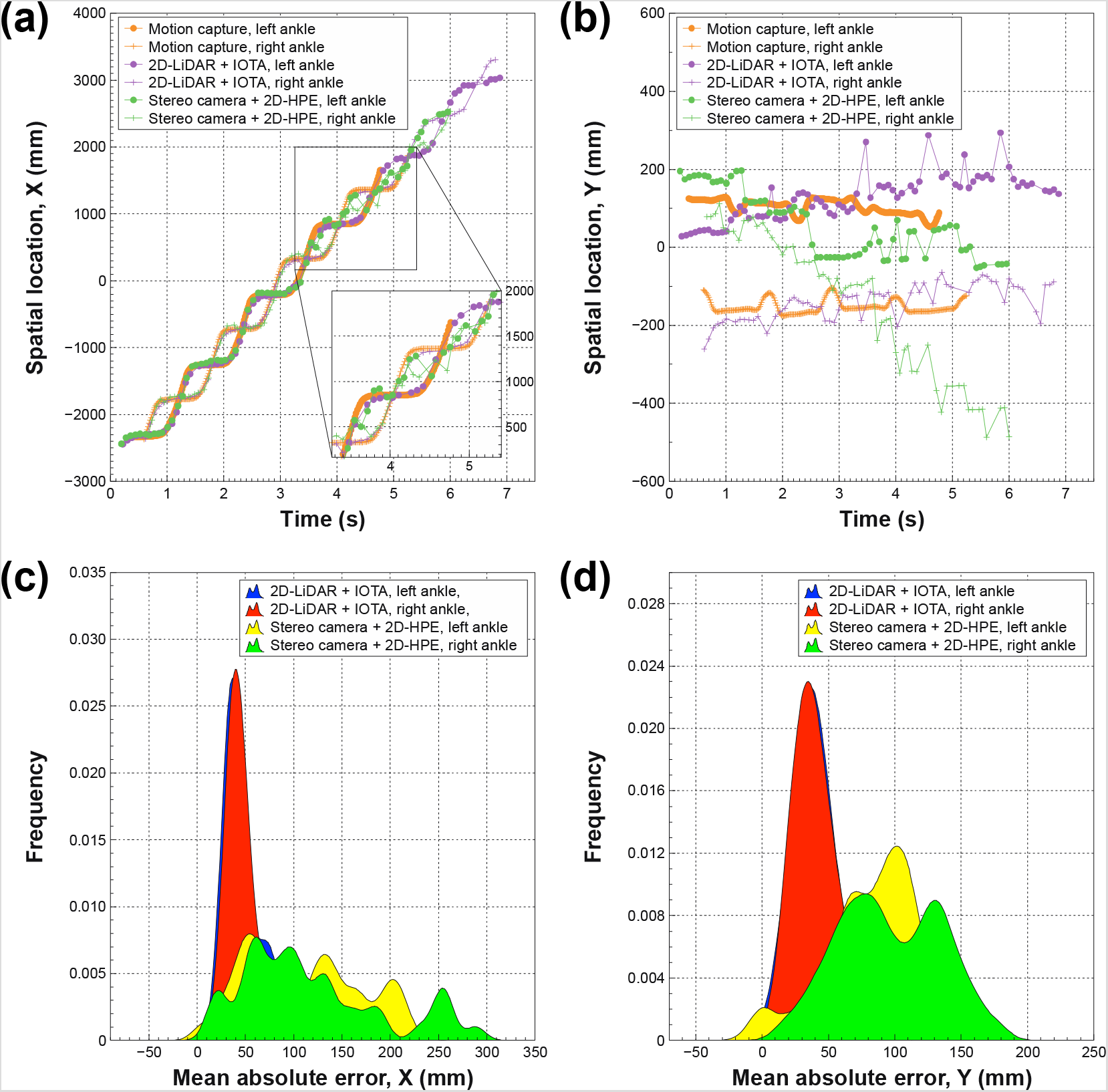
Ankle tracking results and accuracy via 2D-LiDAR with IOTA and stereo camera with 2D-HPE. **a, b** Examples of tracked spatiotemporal locations of ankles along the walking direction (**a**) and the lateral direction **(b)**, gathered by motion capture system, 2D-LiDAR with IOTA, and stereo camera with 2D-HPE. The sensor positions of 2D-LiDAR and stereo camera are at x = –4000 mm. **c, d** Histograms of mean absolute errors of the spatial locations for 2D-LiDAR with IOTA and stereo camera with 2DHPE along the walking direction **(c)** and along the lateral direction **(d)**, respectively, with kernel density estimation smoothing (bandwidth = 10).

Figures 2c and d show the mean absolute error (MAE) of the spatial locations of ankles along the axial and lateral directions to the walking direction, respectively, for 2D-LiDAR with IOTA and stereo camera with 2D-HPE. The average MAEs for left/right ankles on 2D-LiDAR with IOTA and stereo camera with 2D-HPE were 45.1 ± 16.3 mm/47.3 ± 19.4 mm and 114.5 ± 66.3 mm/118.1 ± 73.6 mm, respectively, along the axial direction (Fig. 2c). The low mean values and shallow distributions in MAE on 2D-LiDAR with IOTA are clearly seen. In the lateral direction, the average MAEs were 40.6 ± 16.7 mm/40.6 ± 16.5 mm and 87.8 ± 36.8 mm/96.7 ± 38.8 mm on 2D-LiDAR with IOTA and stereo camera with 2D-HPE, respectively (Fig. 2d).

### Correlations between gait parameters derived from three different modalities

The regressions and Bland-Altman plots for various gait parameters derived from the spatiotemporal locations of ankles on 2D-LiDAR with IOTA and stereo camera with 2D-HPE, where the gait parameters from the motion capture system were regarded as references, are shown in Figs. 3 and 4. Not only the step length, step width, cadence, and gait speed, other spatial and temporal gait parameters was also extracted as shown in supplementary information (Supplementary Figs. 2–4). As shown in Figs. 3a and 4a, strong correlations are seen between 2D-LiDAR with IOTA and motion capture system for step length (r = 0.955, p < 0.001) and cadence (r = 0.911, p < 0.001), while the correlations between stereo camera with 2D-HPE and motion capture system are weaker for step length (r = 0.555, p < 0.001) and cadence (r = 0.510, p < 0.001). The Bland-Altman plot in Fig. 3c and 4c also showed a narrower error distribution for 2D-LiDAR with IOTA than for stereo camera with 2DHPE. For step width, on the other hand, weak correlations are observed for both 2D-LiDAR with IOTA (r = 0.623, p < 0.001) and stereo camera with 2D-HPE (r = –0.098, p = 0.551), as shown in Fig. 3b, since the spatial accuracy along the lateral direction was poor (Fig. 2b). In gait speed, the strongest correlations were observed for both of 2D-LiDAR with IOTA (r = 0.986, p < 0.001) and stereo camera with 2D-HPE (r = 0.903, p < 0.001), as shown in Fig. 4b, where the errors are mostly within 95% confidence intervals (Fig. 3d). It is also noteworthy that the bias in 2D-LiDAR with IOTA was close to zero (Figs. 3c-d and 4c-d), while the bias in stereo camera with 2D-HPE shifted from zero for some cases (Figs. 3d, 4c-d).

**Figure 3.**
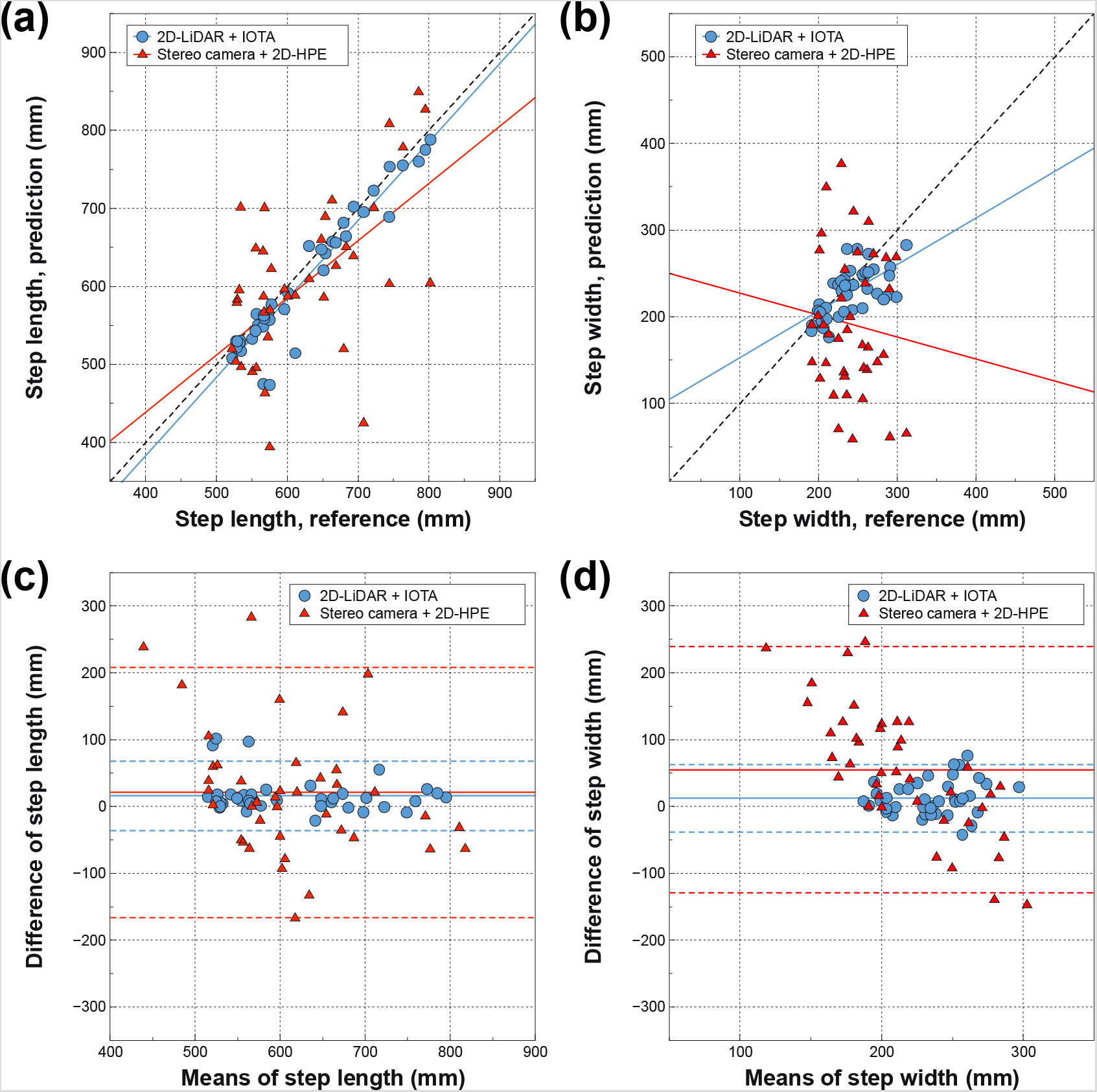
Step length and width derived from spatiotemporal locations of tracked ankles via 2D-LiDAR with IOTA and stereo camera with 2D-HPE. **a-b** Regression and **c-d** Bland-Altman plots with motion capture system for 2D-LiDAR with IOTA and stereo camera with 2D-HPE for step length **(a, c)** and step width **(b, d)**. The black dotted lines in the regression plots are ideal lines for perfect correlations. In the Bland-Altman plots, each solid horizontal line represents the bias of each modality. The upper and lower dashed horizontal lines from each bias represent 95% confidence intervals.

**Figure 4.**
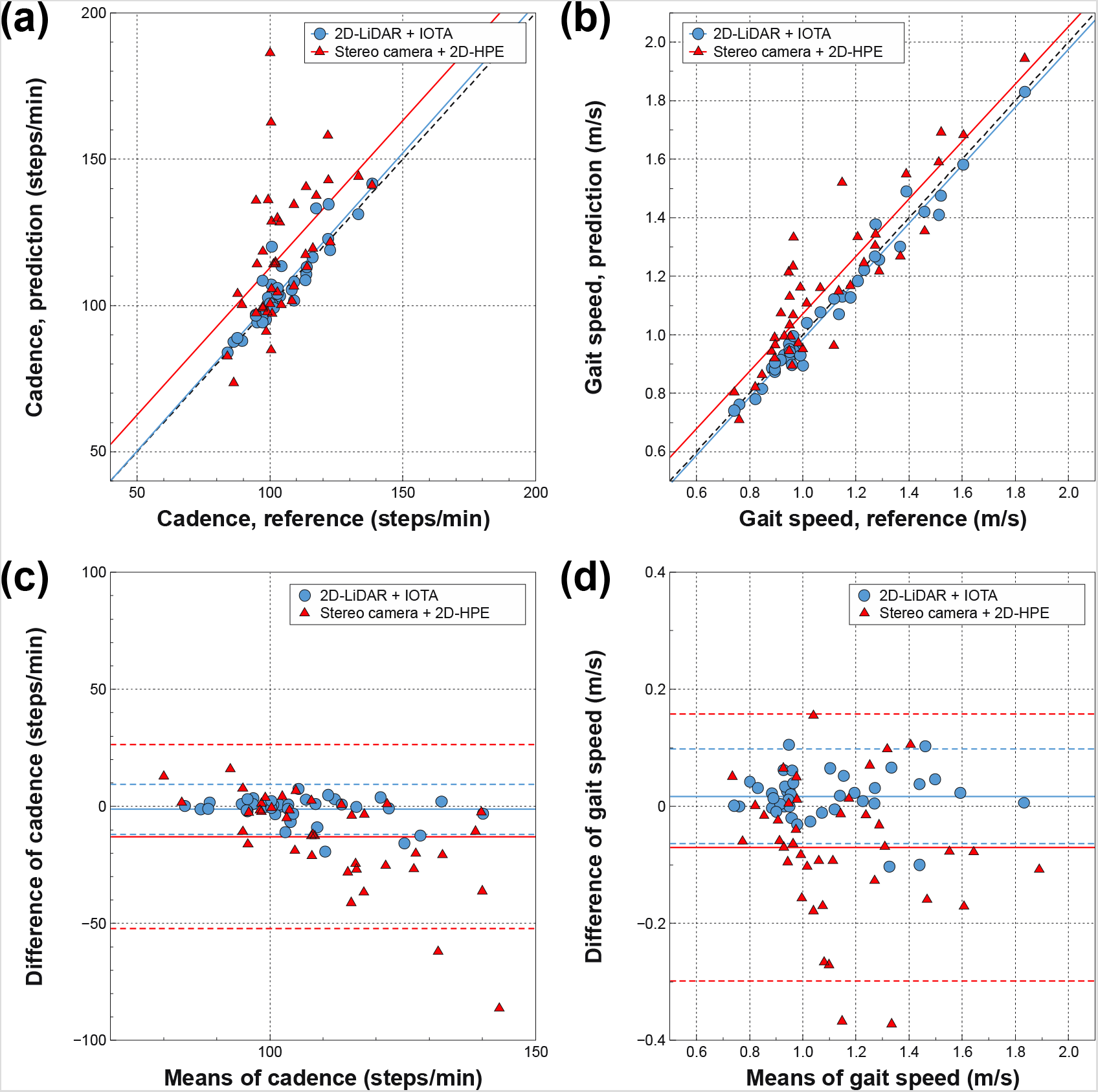
Cadence and gait speed derived from spatiotemporal locations of tracked ankles via 2D-LiDAR with IOTA and stereo camera with 2D-HPE. **a-b** Regression and **c-d** Bland-Altman plots with motion capture system for 2D-LiDAR with IOTA and stereo camera with 2D-HPE for cadence **(a, c)** and gait speed **(b, d)**. The black dotted lines in the regression plots are ideal lines for perfect correlations. In the Bland-Altman plots, each solid horizontal line represents the bias of each modality. The upper and lower dashed horizontal lines from each bias represent 95% confidence intervals.

### Demonstration of multiple target tracking by 2D-LiDAR with IOTA

Since 2D-LiDAR sensor collects spatiotemporal information inherently in a non-intrusive manner, we assessed the expandability of 2D-LiDAR with IOTA to track multiple targets without any instrumental modifications. Figure 5 shows an example of multi-target tracking to mimic a gait monitoring situation where one subject is targeted while two unwanted persons are crossing the monitoring region (see the test environment and walking of subjects in Supplementary Video 2). Before the subject started to walk (t < 2.20 s), the ankles of the subject were still being tracked but not targeted because the left and right ankles was not yet determined (non-targeted object ID 3 and 4 in Fig. 5a). During the initial movement of object snapshots with ID 3 and 4 (i.e., the subject started to walk), the left and right ankles were determined automatically (t < 3.36 s). Meanwhile, the other object snapshots were being tracked concurrently (non-targeted ID 1, 2, 5, and 6 in Fig. 5a). After determining the target ankles, the target object IDs were remembered. During the entire gait monitoring test, a few occlusion events occurred. Figures 4b and c show an example case of the occlusion events, where the object snapshots with ID 1 and 2 had concealed the object snapshots with ID 5 and 6 from the line-of-sight of 2D-LiDAR sensor, which led to missing the spatiotemporal locations of the object snapshots with ID 5 and 6 for some time (∼0.5 s). Because of the occlusion, the tracking became erroneous (object ID 5 in Fig. 5b) or halted (missing traces of the objects ID 5 and 6 in Fig. 5c). Nonetheless, once the object snapshots appeared again, the tracking was successfully restored with appropriate object IDs because IOTA assesses the most probable matching of the object snapshots based on the inertia (Fig. 5c). Similarly, for the targeted ankles (i.e., targeted object ID 3 and 4), two occlusion events were inevitable where the occlusions due to the object snapshots with ID 5 and 6 were the first, followed by object snapshots with ID 1 and 2. Despite the occlusions, tracking of the targeted ankles was well maintained as presented in walking traces in Fig. 5d. A full video clip for the multi-target tracking in Fig. 5 is shown in Supplementary Video 3. Other multi-target tracking tests that simulated various situations, including assisted walk, horizontal walk, and random walk, are also shown in Supplementary Videos 4–6.

**Figure 5.**
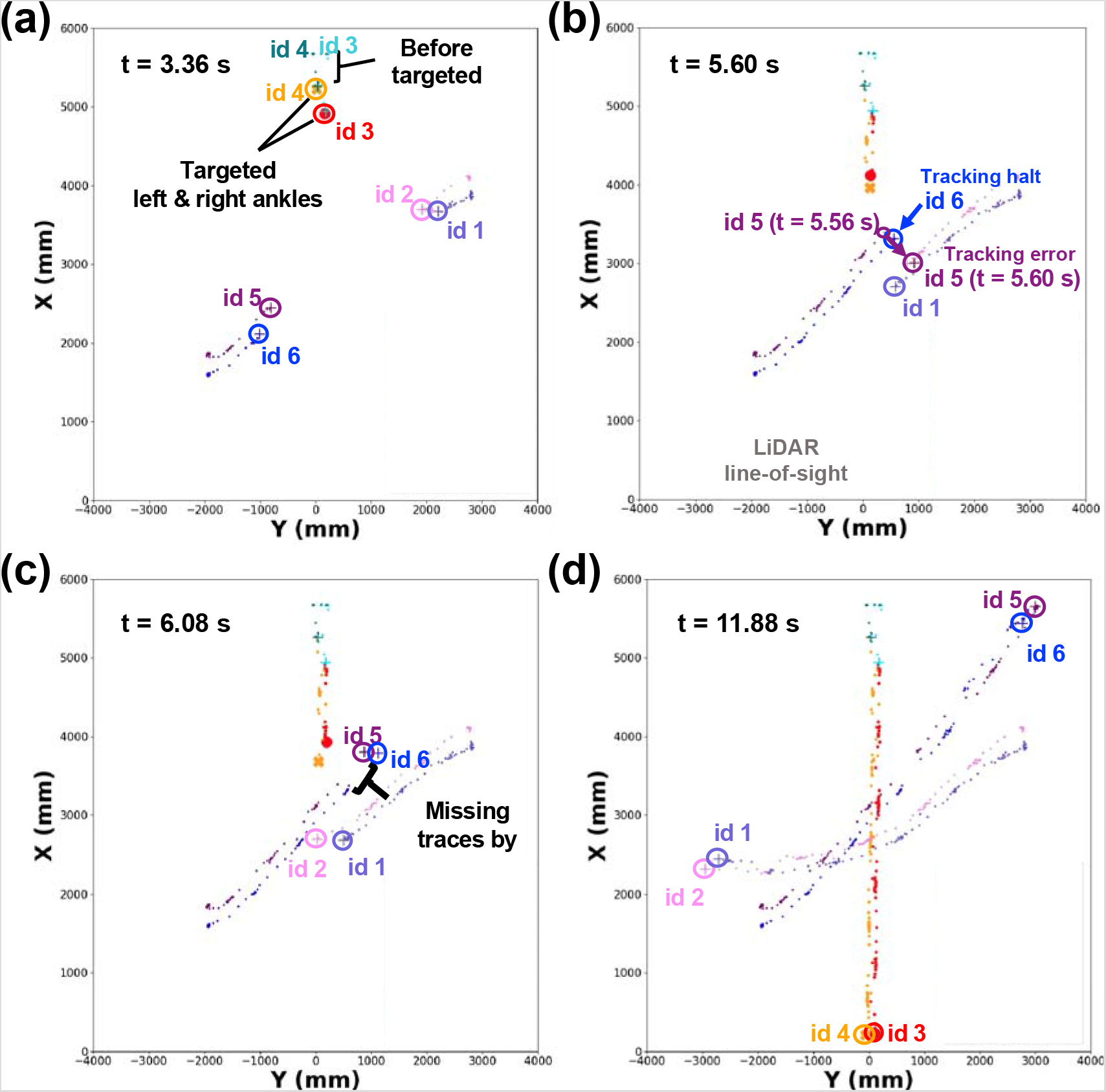
Tracking test for multiple targets via 2D-LiDAR with IOTA. **a-d** Tracked object snapshots for various time frames with the initial detection of the target ankles **(a)**, the occurrence of occlusion **(b)**, the restoration of tracking from the occlusion **(c)**, and the end of measurement **(d)**. Total three persons (i.e., six feet) with the targeted left (ID 3) and right (ID 4) ankles, and untargeted ankles (ID 1, 2, 5, and 6) are presented.

## Discussion

In this study, we established a small-sized, portable 2D-LiDAR-based gait analysis system that can be used in spaces typically up to ∼100 m^2^ to assess gait parameters of multiple persons in a non-intrusive manner and found that the accuracy of parameters from the newly developed system was comparable to that of a dedicated motion capture system. To the authors’ knowledge, this study is the first to use 2D-LiDAR to examine human physical parameters of walking.

Specifically, in our study using 2D-LiDAR with IOTA, the spatial locations along the walking direction corresponded with those of the motion capture system, even though the tracked point data were sparse. Since 2D-LiDAR sensor directly measures absolute distances of objects from the sensor based on a time-of-flight of laser, the measured depth information is highly accurate and reliable. Nevertheless, some inaccuracies in ankle tracking were inevitable at far distances. The main reason for the inaccuracy is a lack of point data at far distances. Because of the limited angular resolution of the 2D-LiDAR sensor, the spatial resolution decreases as the distance from the sensor increases, where we observed that the points almost disappear if the distance between the ankles and the sensor is larger than 10 m.

Similarly, for the stereo camera with 2D-HPE, the tracking accuracy decreases as targets move farther from the sensor. However, the severity and mechanism of decline in accuracy are different in the case of 2D-LiDAR with IOTA. Since a stereo camera estimates the distances of objects from the sensor using the disparity in binocular images, a decline in accuracy is inevitable for far objects because of smaller features and textures of the objects in images. In addition to the intrinsic limitations of a stereo camera, an estimation error from the 2D-HPE network is also crucial for tracking accuracy. As subjects move farther, the number of pixels consisting of the images gradually decreases, i.e., image resolution is reduced, diminishing the essential features for recognizing poses in 2D-HPE network [18]. Consequently, corresponding pixel outputs indicating each keypoint, including left and right ankles, become erroneous.

With supporting clinical evidence, researchers have been keen on measuring gait speed and parameters effectively in clinical practice and in research on older adults. Historically, approaches to acquiring gait parameters have been largely divided under two differing concepts, namely the use of wearable sensors [15, 19] and the use of external devices such as stereo camera, pressure sensors [14], beam breakers [20], and sophisticated motion capture systems [15]. These systems had specific advantages and drawbacks in terms of widespread use in studying the physical performance of older adults. For example, when used appropriately and for a long-term period, wearable devices using inertial measurement units can provide data on physical activity and usual gait speed in life space [19]. However, researches showed that long-term compliance to wearable device is poor in the older population [21]. On the other hand, marker-based systems or walkways can only be used in clinical/ laboratory settings, permitting assessment of gait parameters in a cross-sectional manner. While a single, small-sized stereo camera can capture motion parameters, its short range of reliable measurement precludes acquisition of meaningful lengths of walking as observed in this study.

With characteristic advantages, 2D-LiDAR with IOTA may fill these gaps in existing modalities. As Piau et al. suggested in a recent study, measuring walking speed using home-embedded infrared sensors could help differentiate between eventual fallers and non-fallers [22]. In a living space, 2DLiDAR with IOTA may monitor physical activity and physical performance in real time, with wide detection ranges, in a non-intrusive manner and also in a longitudinal manner. Gait instabilities that may be detected in snapshot measurements may be revealed in the real world at times with cognitive burden such as dual-task situations. In this instance, the relationship between the living space measurement of gait parameters and clinic-based measurement to assess frailty and predict adverse health outcomes might be analogous to the clinic-based 12-lead electrocardiogram and loop recording in approaching syncope [23]. Along with assessing gait parameters *per se*, monitoring physical performance in living space may help detect acute exacerbation, deconditioning, and progression of chronic organ diseases such as chronic obstructive pulmonary disease [24, 25] or congestive heart failure in older adults [26]. Further prospective, real-world studies using 2D-LiDAR with IOTA can be used to assess possible clinical benefits of real-time physical performance monitoring in frail multimorbid older adults.

In clinical practice, access to gait analysis is limited in most settings, by restraints in space and human resources. The new system may be used to acquire gait parameters even when patients enter the clinic room. This newer, 2D system may resolve some drawbacks of previously reported wall-attached infrared beam breakers that could be used to measure gait speeds of patients in outpatients geriatric clinics [20]. While beam breakers cannot distinguish between the patient and caregivers who often accompany outpatients, 2D-LiDAR with IOTA can assess parameters of multiple persons simultaneously, as we showed in this study. As the present study showing improved accessibility to gait parameters in future, the relevance of gait parameters to clinical outcomes in various medical or surgical situations should be assessed.

However, our study had several limitations. Although 2D-LiDAR with IOTA can produce gait parameters comparable to motion capture system, whole-body motion analysis cannot be predicted with the technology because 2D-LiDAR sensor only gathers depth data in a two-dimensional plane. With the continuous scaling in sensor footprint, the angular resolution and price of LiDAR sensors have been improving rapidly; future studies on IOTA with 3D-LiDAR would be interesting. In this preliminary study of the protocol and initial validation, only young and healthy participants were included. However, since frail older people tend to walk slower than younger people, the spatiotemporal reliability of newly developed protocol is not likely to differ substantially when used in older adults.

## Conclusions

The 2D-LiDAR with IOTA was proposed as a practical gait analysis solution for small and noisy clinical environments. Its tracking ability of left and right ankles was validated by comparing it with the motion capture system, which is the gold standard, and with stereo camera with 2D-HPE as a potential alternative. A good agreement between the 2D-LiDAR with IOTA and motion capture system was observed, where the spatial inaccuracy in MAE was as low as 46.2 ± 17.8 mm, which is much better than 116.3 ± 69.6 mm of the stereo camera with 2D-HPE. Based on the advantages of the 2D-LiDAR sensor, such as cost-effectiveness, highly accurate depth measurement, small footprint, and simple installation characteristics, 2D-LiDAR with IOTA can be a promising solution for clinical environments where a simple and quick gait analysis is necessary.

## Data Availability

The datasets generated during the current study are available from the corresponding authors upon reasonable request.

## Acknowledgments

This study was supported by grant No.0420202070 from the Seoul National University Hospital Research Fund.

## Author contributions

S.Y and H.W.J contributed to equally to this work. S.Y, H.W.J, H.J, K.K, S.K.H, H.R, and B.M.O contributed to original research design of study. S.Y, H.W.J, H.J, S.K.H, H.R, and B.M.O contributed to data collection and experimental study. S.K.H contributed to pre- and post-processing for data refinement of motion captured data. S.Y, H.W.J and H.R contributed to development of tracking algorithm for 2D-LiDAR, stereo camera, and analyzed data. S.Y, H.W.J, H.J, K.K, S.K.H, H.R, and B.M.O contributed to paper writing.

## Additional information

**Supplementary information** is available for this paper

**Correspondence** and requests for materials should be addressed to Hyunchul Roh or Byung-Mo Oh

